# Nitroglycerin use preceding primary percutaneous coronary intervention is associated with adverse clinical outcomes in elderly patients with acute coronary syndrome

**DOI:** 10.1101/2023.08.23.23294517

**Authors:** Soichi Komaki, Yunosuke Matsuura, Hiroki Tanaka, Kohei Moribayashi, Yoshimasa Yamamura, Kazumasa Kurogi, Takeshi Ideguchi, Nobuyasu Yamamoto, Michikazu Nakai, Toshihiro Tsuruda, Koichi Kaikita

**Affiliations:** Department of Cardiovascular Medicine, Miyazaki Prefectural Nobeoka Hospital, Nobeoka, Japan; Division of Cardiovascular Medicine and Nephrology, Department of Internal Medicine, Faculty of Medicine, University of Miyazaki, 5200 Kiyotakecho Kihara, Miyazaki-city, Miyazaki 889-1692, Japan; Clinical Research Support Center, University of Miyazaki Hospital, Miyazaki, Japan; and the; Cardiorenal Research Laboratory Department of Hemo-Vascular Advanced Medicine Faculty of Medicine, University of Miyazaki, Miyazaki, Japan

**Keywords:** nitroglycerin, primary care, acute coronary syndrome, percutaneous coronary intervention, elderly patients, major adverse cardiovascular events

## Abstract

**Background:** The primary care for acute coronary syndrome (ACS) includes the administration of nitroglycerin (GTN). This study aimed to investigate the association between the use of GTN before percutaneous coronary intervention (PCI) for ACS and clinical outcomes.

**Methods and Results:** Nine-hundred and forty-seven patients who underwent PCI for ACS were examined and classified into two groups: those who were treated with GTN before PCI (GTN group) and those who were not (non-GTN group). The incidence of major adverse cardiovascular events (MACE), which consist of all-cause mortality, myocardial infarction, stroke, and rehospitalization for heart failure at 1 year, was compared between the two groups. This study identified 289 patients with ACS who used GTN preceding PCI. Pre-PCI systolic blood pressure was significantly lower in the GTN group than in the non-GTN group (median (interquartile range); 130.0 (112.5-144.2) mmHg vs. 142.0 (115.0-160.0) mmHg, respectively, p = 0.03). Multivariate Cox regression analysis indicated that GTN use preceding PCI was an independent determinant for the incidence of MACE (hazard ratio, 1.64; 95% confidence interval, 1.17-2.31; p = 0.004). Overall, the incidence of MACE 1 year after PCI for ACS was significantly higher in the GTN group than in the non-GTN group (log-rank test, p = 0.02); however, this trend was consistently found in elderly patients aged ≥ 75 years (p = 0.002) but not in non-elderly patients aged < 75 years (p = 0.77).

**Conclusions:** GTN use preceding PCI for ACS is associated with lower blood pressure and adverse clinical outcomes in elderly patients.

## Introduction

Primary percutaneous coronary intervention (PCI) (1–3) and optimal medical therapy (OMT), including antithrombotic therapy (4) and lipid-lowering therapy (5), have significantly improved cardiovascular outcomes in patients with acute coronary syndrome (ACS). However, ACS remains a leading cause of death worldwide (6–8). To further improve clinical outcomes, strategies have been implemented to shorten the time from the onset of ACS to the initiation of the diagnostic process or medical intervention, which extend to the prehospital or pre-transport phase (9,10). These management protocols, before primary PCI, include the administration of nitroglycerin (GTN).

GTN has long been incorporated as a first-line drug for ACS (6–8). This is primarily because it decreases venous return, thereby reducing left ventricular filling pressure and blood pressure (11) and, consequently, myocardial oxygen demand (12). However, limited studies (13,14) support the use of GTN for improving ACS outcomes; moreover, these investigations were mainly conducted over three decades ago when the clinical demographics of patients with ACS significantly differed from the current situation. The changes in patient characteristics (e.g., an increased number of elderly patients) and advances in ACS management, including PCI and OMT, have raised the need to reassess the efficacy of GTN for clinical outcomes after primary PCI for ACS. Therefore, this study investigated the association between pre-PCI use of GTN and clinical outcomes in patients with ACS.

## Methods

### Study Design and Population

We conducted a single-center, retrospective, observational study of consecutive patients with ACS admitted to Miyazaki Prefectural Nobeoka Hospital from January 2013 to May 2021. Initially, 1131 patients were enrolled. Subsequently, patients who did not undergo revascularization (n=164) and patients who underwent emergency coronary artery bypass surgery (n=20) were excluded, resulting in a total of 947 patients for the final analysis (**Figure 1**, Supplementary Methods S1). The use of GTN preceding primary PCI was defined as the administration of GTN in primary care before or after arrival at the emergency department (ED), except for GTN administration in the cardiac catheterization laboratory. A review of medical records on drug use before and after arrival at our ED confirmed the following: (1) 289 patients received sublingual, oral spray, or IV administration of GTN before PCI (250 patients before transport to our institute and 39 patients due to immediate care after arrival at the ED); (2) 144 patients received a loading dose of antiplatelet agents at the prior hospital; and (3) 45 patients received unfractionated heparin at the prior hospital. In this study, patients with ACS who underwent PCI were divided into the GTN group and the non-GTN group according to the use of GTN before PCI (Supplementary Methods S2). The association between the use of GTN before PCI and clinical outcomes was examined. All procedures included in this study complied with the principles outlined in the Declaration of Helsinki. The study was reviewed and approved by the Institutional Review Board and the Ethics Committee of Miyazaki Prefectural Nobeoka Hospital (Reference Number: 20191004-1) and the University of Miyazaki (Reference Number: O-1277). The need for obtaining informed consent was waived owing to the low-risk nature of the study. This study was disclosed and announced on the hospital website (https://nobeoka-kenbyo.jp/sinryoka/junkankika/) in case any patient wished to opt out.

**Figure 1.**
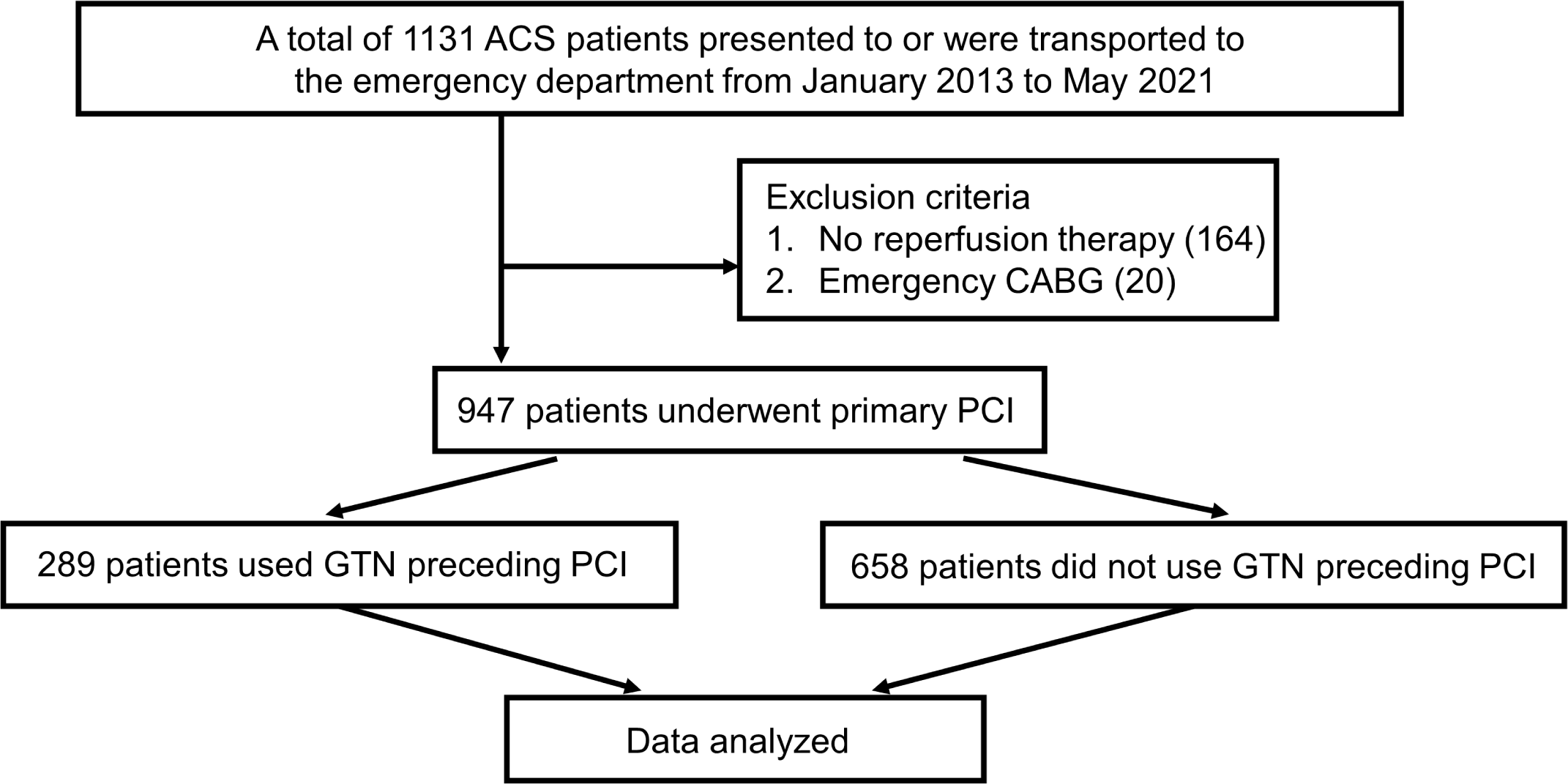
Flow chart of patient inclusion and exclusion. ACS, acute coronary syndrome; CABG, coronary artery bypass grafting; GTN, nitroglycerin; PCI, percutaneous coronary intervention.

### PCI Procedures and Pre- and Post-PCI Medical Management

Based on the relevant ACS treatment guidelines (6), all patients received aspirin, a P2Y12 inhibitor (clopidogrel or prasugrel), and weight-adjusted intravenous heparin before primary PCI (**Supplementary Methods S3**). Coronary angiography and subsequent PCI were performed according to standard practice, and the culprit lesions requiring urgent revascularization were determined by angiography in conjunction with electrocardiographic and echocardiographic findings. The door-to-balloon (D2B) time was measured from admission to achieving coronary reperfusion, and TIMI grades were evaluated before and after PCI. Successful reperfusion was defined as the confirmation of TIMI 3 flow on angiography. The indications for emergency coronary artery bypass grafting (CABG) included the following: (1) the presence of active ischemia with contraindications for PCI; (2) successful PCI of the culprit lesion and further indication for CABG; and (3) incomplete, insufficient, or failed PCI. Patients with hemodynamic compromise refractory to medical treatment and revascularization for ACS were treated with cardiac and respiratory assist devices, including intra-aortic balloon pumping, veno-arterial external extracorporeal membrane oxygenation, temporary pacing, non-invasive positive pressure ventilation, and mechanical ventilation with endotracheal intubation, as appropriate.

All patients who had undergone primary PCI for ACS immediately received comprehensive treatment, which included the timely initiation of OMT in the coronary care unit, where cardiac biomarkers, including creatine kinase (CK), were measured and monitored.

Additionally, cardiac rehabilitation was initiated at the appropriate time for the peak CK level according to standard practice.

### Clinical Outcomes

Clinical follow-up information was obtained from medical records and/or telephone interviews with patients or their families. The primary outcome was the incidence of major adverse cardiovascular events (MACE) within 1 year, defined as a composite of all-cause death, non-fatal myocardial infarction (MI), stroke, and rehospitalization for heart failure. The secondary outcome was target lesion revascularization (TLR) after stent implantation at the 1-year follow-up. TLR was defined according to the Academic Research Consortium criteria (15).

### Statistical Analysis

Categorical and continuous variables are presented as numbers with percentages and as median values with interquartile ranges, respectively. The chi-square test or Fisher’s exact test was used for categorical variables, and the Mann-Whitney U test was used for continuous variables. In the GTN and non-GTN groups, clinical variables such as patient characteristics (including comorbidities, history, laboratory data, medication, and PCI procedure relevant factors) and clinical outcomes were compared. The cumulative incidence rates of MACE were calculated by the Kaplan-Meier method, with comparisons between the GTN and non-GTN groups being performed using log-rank tests. Univariate and multivariate Cox regression analyses were used to calculate hazard ratios (HRs) and corresponding 95% confidence intervals (CIs) for the incidence of MACE. Multivariate analyses were performed using statistically significant clinical variables from the univariate analysis. In addition, inverse probability weighting (IPW) was performed as a sensitivity analysis using the same covariates with multivariate analysis. Statistical significance was defined as a p-value of less than 0.05, and all statistical analyses were performed using SPSS version 20.0 (IBM Corp., Armonk, NY, USA) and STATA 18 (College Station, TX, USA).

## Results

### Clinical Characteristics in the GTN and the Non-GTN Groups

This study analyzed 947 patients with a median age of 71 years and with 71.5% of male patients. Hypertension was the most common coronary risk factor, followed by dyslipidemia, smoking, and diabetes. Two hundred eighty-nine patients (30.5%) received GTN preceding PCI as primary care for ACS in prehospital and in-hospital phases: 228 with sublingual or oral spray, 57 with intravenous, and 4 with both. In the pre-PCI baseline characteristics, the GTN group (vs. non-GTN group) had significantly lower systolic blood pressure [130 (112–144) mmHg vs. 142 (115–160) mmHg, p = 0.03], a trend toward lower diastolic blood pressure [82 (68–95) mmHg vs. 84 (70–96) mmHg, p = 0.08], and significantly higher prevalence of hypertension (82.6% vs. 72.6%, p < 0.001) and dyslipidemia (76.8% vs. 66.1%, p < 0.001). In addition, the GTN group had a trend toward a higher proportion of patients who previously received PCI (17.9% vs. 13.3%, p = 0.065) (Supplementary Table S1). In a survey of antithrombotic pharmacotherapy regimens before the onset of ACS, the GTN group showed significantly higher rates of aspirin (20.7% vs. 12.7%, p = 0.003) and warfarin use (2.4 % vs. 0.5%, p = 0.016) and a trend toward higher rates of direct oral anticoagulant (DOAC) use (2.4% vs. 0.8%, p = 0.066), compared with the non-GTN group (Supplementary Table S1).

No significant differences were found between the GTN and non-GTN groups regarding PCI approach site, target vessel, Killip classification, frequency of drug-eluting stent and bare metal stent use, peak level of CK, D2B time, and frequency of imaging device use, except for that the GTN group showed a significantly lower proportion of RCA infarction and temporary pacing therapy than the non-GTN group in emergency PCI procedures (Supplementary Table S2). Regarding antithrombotic drugs at discharge, 98.9% of the patients received aspirin, 38.9% received clopidogrel, 59.6% received prasugrel, 2.3% received warfarin, and 6.2% received DOACs. No difference in the proportion of antithrombotic drugs was observed between the GTN and non-GTN groups (Supplementary Table S2).

### Clinical Outcomes in the GTN and Non-GTN Groups

PCI was successfully performed for 942 (99.4%) patients. During the 1-year follow-up, the incidence of MACE was found in 146 (15.4%) patients: 88 (9.2%) with all-cause death, 13 (1.3%) with MI, 23 (2.4%) with stroke, and 27 (2.8%) with heart failure requiring rehospitalization. Compared with the non-GTN group, the GTN group exhibited a significantly higher incidence of MACE (19.3 % vs. 13.6%, p = 0.025). In addition, a trend toward the higher incidence of TLR (7.2% vs. 4.5%, p = 0.089) was observed in the GTN than in the non-GTN group (**Table 1**). In the GTN group, patients with MACE had significantly lower levels of systolic [130.0 (95.0-141.0) vs. 132.0 (113.0-146.0) mmHg, p = 0.011] and diastolic blood pressure [76.5 (57.7-91.2) vs. 84.0 (70.0-95.0) mmHg, p = 0.011] than those without MACE (**Supplementary Table S3**).

**Table 1.**
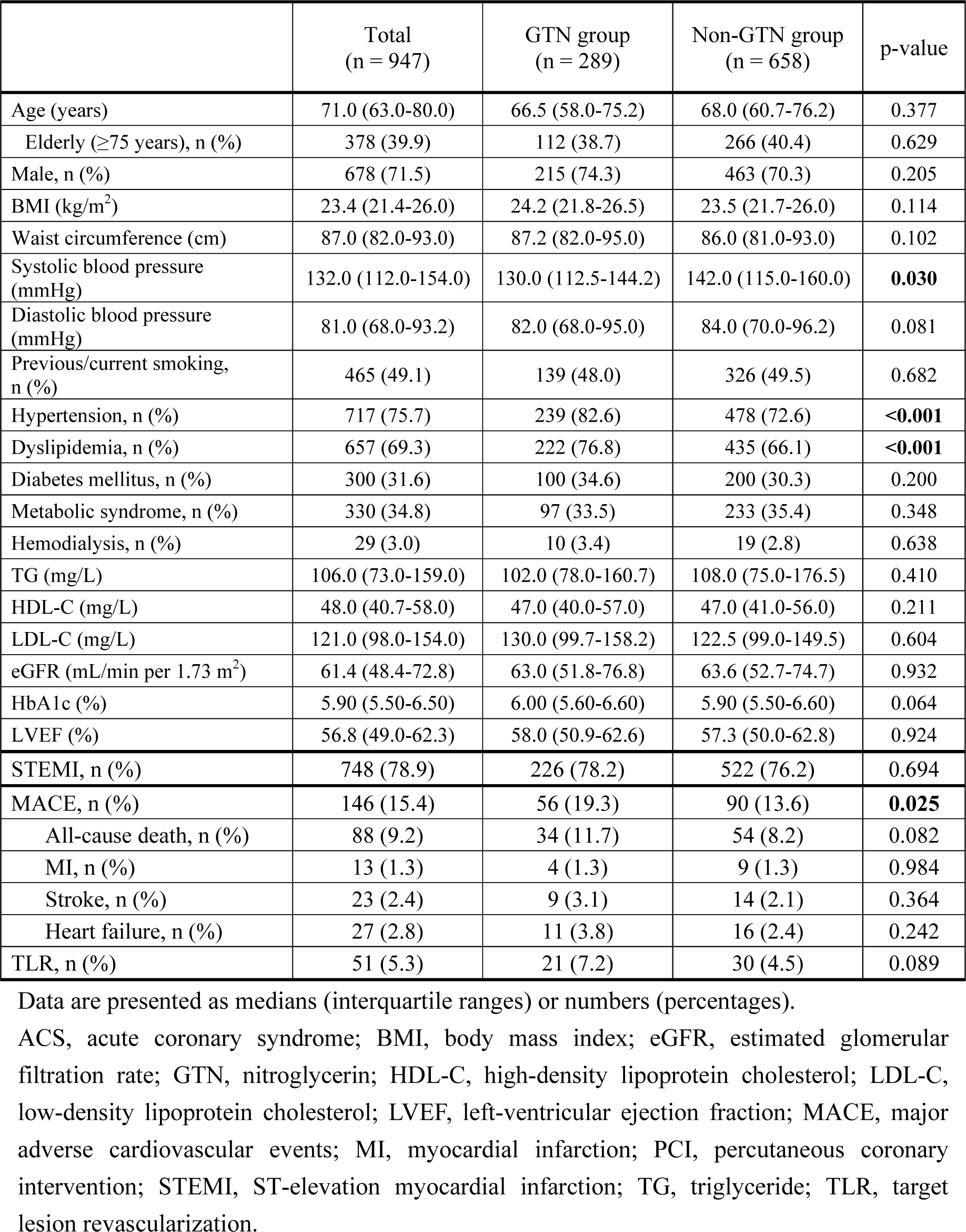
Demographics and the use of GTN before PCI in patients with ACS.

|  | Total<br>(n = 947) | GTN group<br>(n = 289) | Non-GTN group<br>(n = 658) | p-value |
| --- | --- | --- | --- | --- |
| Age (years) | 71.0 (63.0-80.0) | 66.5 (58.0-75.2) | 68.0 (60.7-76.2) | 0.377 |
| Elderly ($\geq 75$ years), n (%) | 378 (39.9) | 112 (38.7) | 266 (40.4) | 0.629 |
| Male, n (%) | 678 (71.5) | 215 (74.3) | 463 (70.3) | 0.205 |
| BMI (kg/m<sup>2</sup>) | 23.4 (21.4-26.0) | 24.2 (21.8-26.5) | 23.5 (21.7-26.0) | 0.114 |
| Waist circumference (cm) | 87.0 (82.0-93.0) | 87.2 (82.0-95.0) | 86.0 (81.0-93.0) | 0.102 |
| Systolic blood pressure (mmHg) | 132.0 (112.0-154.0) | 130.0 (112.5-144.2) | 142.0 (115.0-160.0) | **0.030** |
| Diastolic blood pressure (mmHg) | 81.0 (68.0-93.2) | 82.0 (68.0-95.0) | 84.0 (70.0-96.2) | 0.081 |
| Previous/current smoking, n (%) | 465 (49.1) | 139 (48.0) | 326 (49.5) | 0.682 |
| Hypertension, n (%) | 717 (75.7) | 239 (82.6) | 478 (72.6) | **<0.001** |
| Dyslipidemia, n (%) | 657 (69.3) | 222 (76.8) | 435 (66.1) | **<0.001** |
| Diabetes mellitus, n (%) | 300 (31.6) | 100 (34.6) | 200 (30.3) | 0.200 |
| Metabolic syndrome, n (%) | 330 (34.8) | 97 (33.5) | 233 (35.4) | 0.348 |
| Hemodialysis, n (%) | 29 (3.0) | 10 (3.4) | 19 (2.8) | 0.638 |
| TG (mg/L) | 106.0 (73.0-159.0) | 102.0 (78.0-160.7) | 108.0 (75.0-176.5) | 0.410 |
| HDL-C (mg/L) | 48.0 (40.7-58.0) | 47.0 (40.0-57.0) | 47.0 (41.0-56.0) | 0.211 |
| LDL-C (mg/L) | 121.0 (98.0-154.0) | 130.0 (99.7-158.2) | 122.5 (99.0-149.5) | 0.604 |
| eGFR (mL/min per 1.73 m<sup>2</sup>) | 61.4 (48.4-72.8) | 63.0 (51.8-76.8) | 63.6 (52.7-74.7) | 0.932 |
| HbA1c (%) | 5.90 (5.50-6.50) | 6.00 (5.60-6.60) | 5.90 (5.50-6.60) | 0.064 |
| LVEF (%) | 56.8 (49.0-62.3) | 58.0 (50.9-62.6) | 57.3 (50.0-62.8) | 0.924 |
| STEMI, n (%) | 748 (78.9) | 226 (78.2) | 522 (76.2) | 0.694 |
| MACE, n (%) | 146 (15.4) | 56 (19.3) | 90 (13.6) | **0.025** |
| All-cause death, n (%) | 88 (9.2) | 34 (11.7) | 54 (8.2) | 0.082 |
| MI, n (%) | 13 (1.3) | 4 (1.3) | 9 (1.3) | 0.984 |
| Stroke, n (%) | 23 (2.4) | 9 (3.1) | 14 (2.1) | 0.364 |
| Heart failure, n (%) | 27 (2.8) | 11 (3.8) | 16 (2.4) | 0.242 |
| TLR, n (%) | 51 (5.3) | 21 (7.2) | 30 (4.5) | 0.089 |

Data are presented as medians (interquartile ranges) or numbers (percentages).
ACS, acute coronary syndrome; BMI, body mass index; eGFR, estimated glomerular filtration rate; GTN, nitroglycerin; HDL-C, high-density lipoprotein cholesterol; LDL-C, low-density lipoprotein cholesterol; LVEF, left-ventricular ejection fraction; MACE, major adverse cardiovascular events; MI, myocardial infarction; PCI, percutaneous coronary intervention; STEMI, ST-elevation myocardial infarction; TG, triglyceride; TLR, target lesion revascularization.

**Table 2** shows the results of univariate and multivariate analyses for clinical variables associated with MACE that occurred during follow-up in all patients. Univariate analysis indicated that age, dyslipidemia, chronic kidney disease, Killip classification, and the use of GTN were significantly associated with the incidence of MACE (all, p <0.05). Multivariate analysis indicated that in addition to dyslipidemia, chronic kidney disease, and Killip classification (all, p < 0.05), the use of GTN was an independent determinant for the incidence of MACE (HR, 1.64; 95% CI, 1.17-2.31; p = 0.004).

**Table 2.** Univariate and multivariate analysis and inverse probability weighting on the predictor for MACE after PCI.

|  | Univariate analysis |  |  | Multivariate analysis |  |  | Inverse probability weighting |  |  |
| --- | --- | --- | --- | --- | --- | --- | --- | --- | --- |
|  | HR | 95% CI | p-value | HR | 95% CI | p-value | HR | 95% CI | p-value |
| Age | 1.02 | 1.01-1.04 | **0.001** | 1.00 | 0.99-1.02 | 0.619 |  |  |  |
| Male | 0.76 | 0.54-1.07 | 0.117 |  |  |  |  |  |  |
| BMI | 0.97 | 0.92-1.02 | 0.263 |  |  |  |  |  |  |
| Hypertension | 1.08 | 0.74-1.60 | 0.680 |  |  |  |  |  |  |
| Dyslipidemia | 0.61 | 0.44-0.84 | **0.003** | 0.61 | 0.44-0.86 | **0.005** |  |  |  |
| Diabetes mellitus | 1.21 | 0.86-1.70 | 0.274 |  |  |  |  |  |  |
| Chronic kidney disease | 2.68 | 1.89-3.79 | **<0.001** | 2.07 | 1.43-3.01 | **<0.001** |  |  |  |
| Killip classification | 4.40 | 3.09-6.25 | **<0.001** | 3.78 | 2.64-5.40 | **<0.001** |  |  |  |
| STEMI | 0.81 | 0.56-1.19 | 0.286 |  |  |  |  |  |  |
| Prior stroke | 1.41 | 0.86-2.30 | 0.176 |  |  |  |  |  |  |
| GTN use before PCI | 1.46 | 1.05-2.04 | **0.026** | 1.64 | 1.17-2.31 | **0.004** | 1.56 | 1.14-2.13 | **0.005** |

BMI, body mass index; CI, confidence interval; eGFR, estimated glomerular filtration rate; GTN, nitroglycerin; HR, hazard ratio; LVEF, left-ventricular ejection fraction; MACE, major adverse cardiovascular events; PCI, percutaneous coronary intervention; STEMI, ST-elevation myocardial infarction.
Age; 1-y increase, BMI categories 18.5-26, $\leq 18.5$ or $> 26$ ; LVEF $\geq 50\%$ , $< 50\%$ ; chronic kidney disease = eGFR $\geq 60$ mL/min per $1.73 \text{ m}^2$ , eGFR $< 60$ mL/min per $1.73 \text{ m}^2$ ; Killip classification 1 or 2, 3, 4.

As shown in **Figure 2A**, Kaplan-Meier curves in all patients revealed that the incidence of MACE was significantly higher in the GTN group than in the non-GTN group (p = 0.024). Next, the difference in MACE incidence between the GTN and non-GTN groups was analyzed separately for elderly (age ≥75 years) and non-elderly patients (age <75 years); the significantly higher incidence of MACE in the GTN group than the non-GTN group was found only in elderly patients (p = 0.002) (**Figure 2B**) and not in non-elderly patients (p = 0.773) (**Figure 2C**).

**Figure 2.**
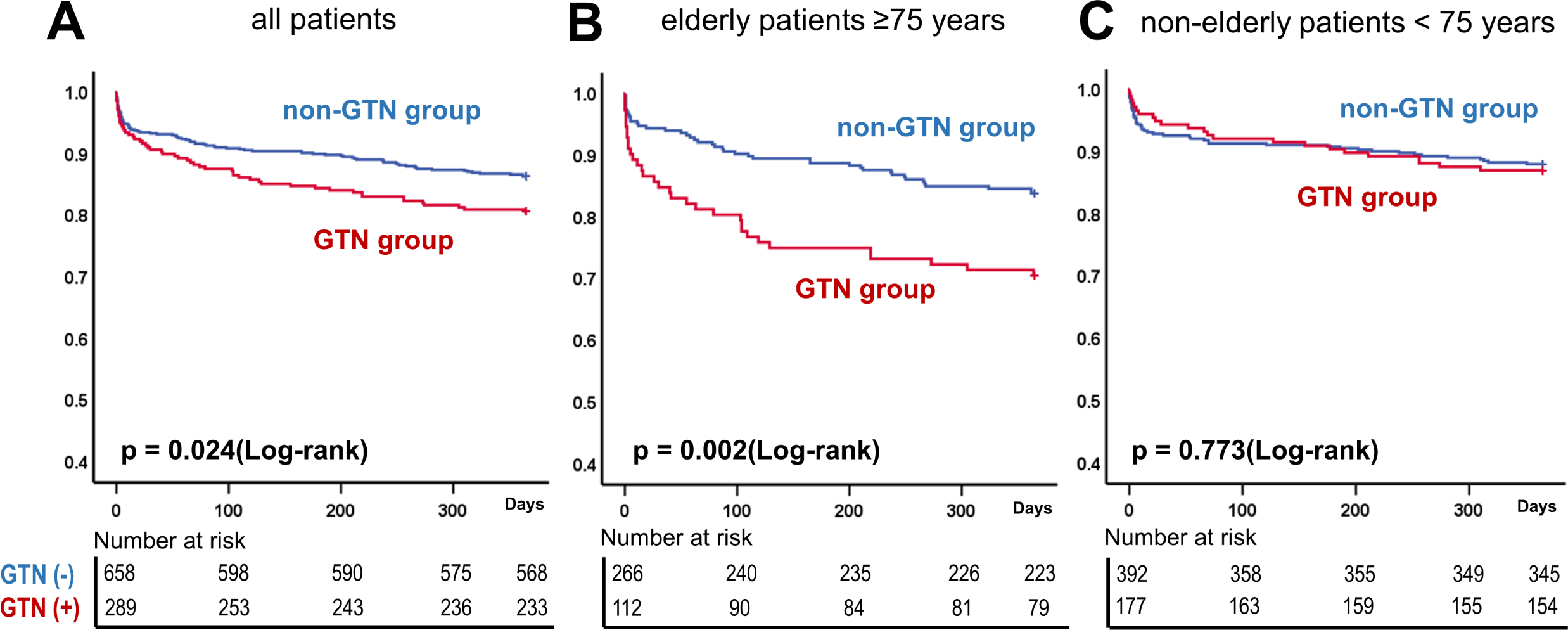
Comparison of the incidence of MACE using Kaplan-Meier Curves between the GTN and non-GTN groups in all patients, elderly, and non-elderly patients with ACS. **(A)** patients of all ages **(B)** elderly patients (≥75 years) **(C)** non-elderly patients (< 75 years) ACS, acute coronary syndrome; GTN, nitroglycerin; MACE, major adverse cardiac events.

Pre-PCI baseline characteristics of the elderly patients indicated that the GTN group had a significantly lower systolic blood pressure [132.0 (101.7-141.0) vs. 139.0 (112.0-155.0) mmHg, p = 0.007] and higher prevalence of dyslipidemia (76.7% vs. 60.1%, p = 0.002) than the non-GTN group. In addition, the GTN group in elderly patients tended to receive aspirin before ACS (25.0% vs. 16.5%, p = 0.056) (**Supplementary Table S4**). Multivariate analysis in elderly patients indicated that Killip classification and the use of GTN were independent determinants for the incidence of MACE (HR, 2.33; 95% CI, 1.44-3.75; p <0.001, HR, 1.89; 95% CI, 1.19-2.97; p = 0.006) (**Table 3**). As a sensitivity analysis, IPW showed similar results with multivariate analysis (**Tables 2 and 3**).

**Table 3.**
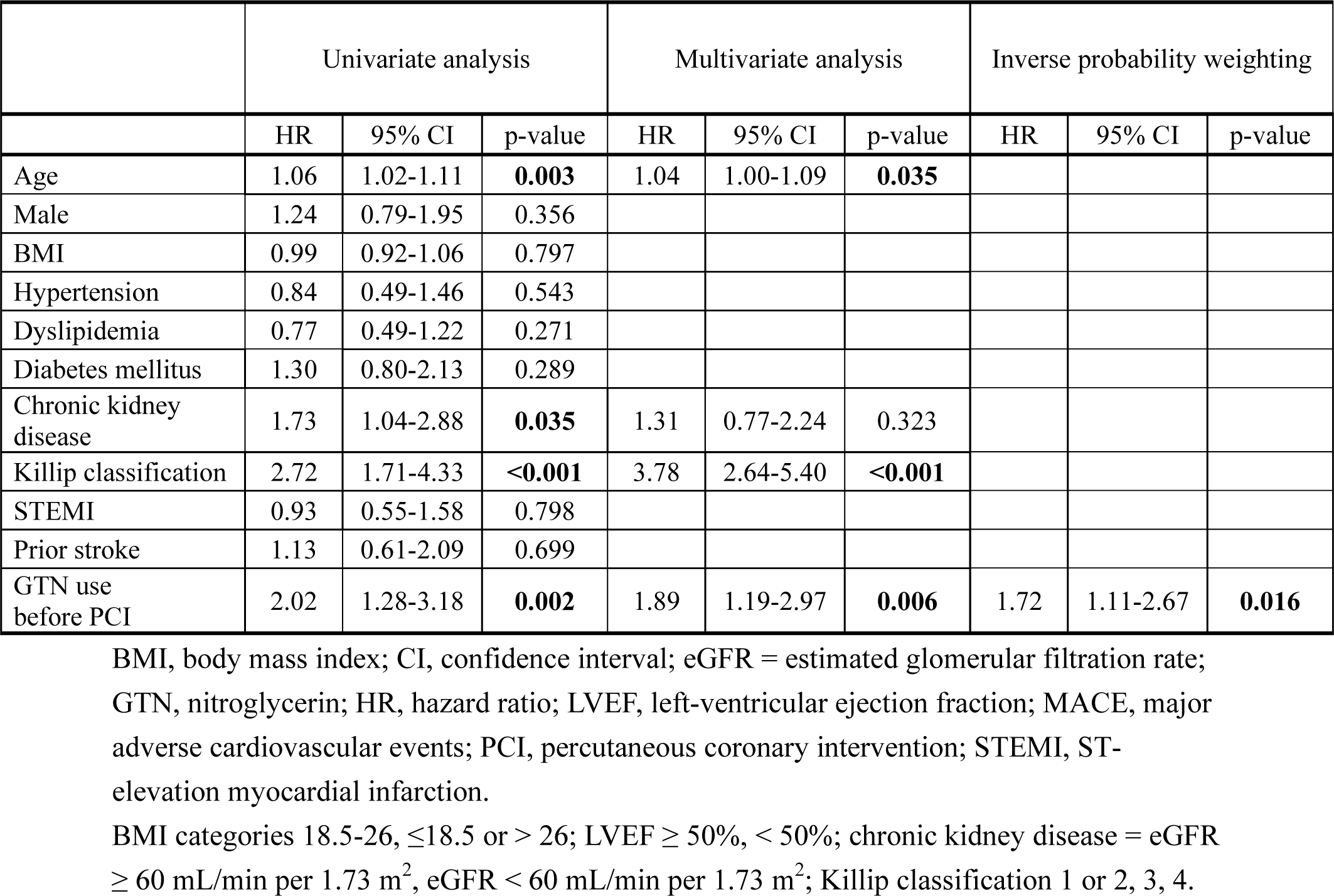
Univariate and multivariate analysis and inverse probability weighting on the predictor for MACE after PCI in elderly patients (≥ 75 years) with ACS.

## Discussion

This study investigated the association between GTN use preceding PCI for ACS and the incidence of MACE. The noteworthy findings are as follows: 1) Overall, the GTN group had lower pre-PCI blood pressure and a significantly higher incidence of MACE at 1-year follow-up the non-GTN group, although peak CK level and left-ventricular ejection fraction (LVEF) after PCI for ACS were similar in the two groups. In addition to dyslipidemia, chronic kidney disease, and Killip classification, GTN use was an independent determinant for the incidence of MACE. 2) The GTN group had a significantly higher incidence of MACE than the non-GTN group in elderly patients (age ≥75 years); however, the incidence of MACE did not differ between the GTN and non-GTN groups in non-elderly patients (age <75 years). 3) In both the GTN and non-GTN groups, patients who developed MACE had lower pre-PCI blood pressure than those who did not develop MACE. Moreover, the GTN group showed less pre-PCI systolic blood pressure than the non-GTN group among elderly patients, but we did not find such a difference between the GTN and non-GTN groups in non-elderly patients.

The latest guidelines from the Japanese Circulation Society (6), and American College of Cardiology/American Heart Association (8) recommend the administration of nitrates as a Class I indication in the primary management for ACS, except in cases with contraindications such as marked hypotension, bradycardia, and complicated right ventricular infarction.

However, as Ekmejian et al. recently described (16), these guidelines rely on the results from the GISSI-3 (13) and ISIS-4 study (14) conducted more than 30 years ago when the treatment of ACS and the demographics of patients with ACS were significantly different from the current situation. Moreover, the results of these studies did not fully support the strong recommendations for the use of GTN for ACS. The percentage of patients with ACS undergoing primary PCI followed by OMT has increased markedly over the past 30 years, and these multidisciplinary treatments have significantly suppressed the development and progression of heart failure and the recurrence of ischemic events. Additionally, the recent promotion of timely reperfusion after the onset of ST-elevation myocardial infarction includes shortening the D2B time to less than 90 minutes (17,18). Our hospital data also indicated that the D2B time was 73 minutes, meeting the goal stated in the guidelines (6). All of these initiatives help preserve cardiac function after ACS, resulting in the hemodynamically beneficial effect of GTN in the acute phase, which might have become relatively limited and less apparent compared with the era without widespread PCI and OMT. The concerns regarding the safety of GTN use for ACS might have been relatively increased because the number and proportion of elderly patients with ACS increased, and elderly patients are more prone to hypotension after GTN administration (19). The present study also confirmed that the GTN group showed less pre-PCI systolic blood pressure than the non-GTN group among elderly patients. Elderly patients 75 or more years of age account for 30-40% of all hospitalized patients with ACS, and the incidence of ACS-associated death is also mainly observed in this age group (20–22). Similarly, the present study included approximately 40% of patients aged 75 or more years, and the trend of outcomes for the study population as a whole and those in this age group were nearly identical. These findings suggest that the incidence of clinical events occur mainly in elderly patients, which is similar to previous reports. Hence, optimization of primary management for elderly patients with ACS is required (23), and the pros and cons of GTN use should be debated and examined separately for elderly and non-elderly patients.

The in-hospital and 1-year mortality among patients with ACS can be effectively estimated and identified by the GRACE score (24,25), which consists of eight items, including age and systolic blood pressure. The results of our study indicated a higher incidence of MACE in the GTN rather than the non-GTN group among elderly patients but not among non-elderly patients. Although this study lacks the GRACE score, poor outcomes in the elderly GTN group might be explained partly by risk elevation owing to the accumulation of older age and less systolic blood pressure. A future study on whether the risk stratification score incorporating ‘‘GTN use before primary PCI’’ can be developed in patients with ACS may be required.

### Limitations

This study had several limitations. First, the study was a single-center cohort. Since the PCI strategy directly influences the incidence of MACE, generalizing these single-center results beyond strategical differences requires multicenter validation. Second, the results do not allow us to simply conclude that GTN use preceding primary PCI causally decreases blood pressure on admission and worsens clinical outcomes; prospective interventional studies are warranted. Third, the present study failed to show data on the time from the onset of chest symptoms to the use of GTN. Although there was no difference in peak CK elevation or LVEF between the GTN and non-GTN groups, the treatment, including GTN administration itself, might have prolonged the time until primary PCI from symptom onset. Finally, patients with ACS presenting with severer symptoms of chest pain might be more likely to be treated with GTN, suggesting that GTN use may be a marker for more severe ischemia or more compromised patients.

## Conclusions

In this study, GTN use preceding primary PCI was associated with adverse clinical outcomes in elderly patients with ACS. Further studies are needed to re-evaluate the impact of GTN use on clinical outcomes in these patients.

## Clinical Perspective

### What Is New?

Nitroglycerin (GTN) use preceding primary percutaneous coronary intervention (PCI) for acute coronary syndrome (ACS) was an independent determinant for the incidence of the 1-year major adverse cardiovascular events (MACE).

In particular, the GTN group had a significantly higher incidence of MACE than the non-GTN group in elderly patients (age ≥75 years).

### What Are the Clinical Implications?

GTN use preceding primary PCI was associated with adverse clinical outcomes in elderly patients with ACS.

In the optimization of primary management for patients with ACS, the pros and cons of GTN use might need to be debated and examined separately for elderly and non-elderly patients.

### Impact on daily practice

The association between the use of nitroglycerin (GTN) before percutaneous coronary intervention (PCI) for acute coronary syndrome (ACS) and clinical outcomes remains unclear. We conducted a retrospective observational study including 947 ACS patients with or without GTN use preceding primary PCI. The outcome was the 1-year major adverse cardiovascular events (MACE). The incidence of MACE was significantly higher in the GTN group than in the non-GTN group; this trend was consistently found in elderly patients aged ≥ 75 years (p = 0.02 and 0.002, respectively). GTN use preceding PCI for ACS is associated with adverse clinical outcomes in elderly patients.

### Data Availability Statement

Raw data were generated at University of Miyazaki. Derived data supporting the findings of this study are available from the corresponding author on request. The data are not publicly available due to privacy reasons.

**Sources of Funding**: This study was supported by a Grant for Clinical Research from Miyazaki University Hospital.

**Disclosures:** Dr. Koichi Kaikita has received remuneration for lectures from Bayer Yakuhin Ltd., Daiichi-Sankyo Co. Ltd., Novartis Pharma AG., and Otsuka Pharmaceutical Co. Ltd.; has received trust research/joint research funds from Bayer Yakuhin Ltd. and Daiichi-Sankyo Co. Ltd.; and has received scholarship funds from Abbott Medical Co. Ltd. The remaining authors have nothing to disclose.

## Abbreviations

ACS: acute coronary syndrome
CABG: coronary artery bypass grafting
D2B: door-to-balloon
GTN: nitroglycerin
LVEF: left-ventricular ejection fraction
MACE: major adverse cardiovascular events
MI: myocardial infarction
OMT: optimal medical therapy
PCI: percutaneous coronary intervention
TLR: target lesion revascularization

